# Severe COVID-19 infection is associated with increased antibody-mediated platelet apoptosis

**DOI:** 10.1101/2020.09.03.20187286

**Authors:** Karina Althaus, Irene Marini, Jan Zlamal, Lisann Pelzl, Helene Häberle, Martin Mehrländer, Stefanie Hammer, Harald Schulze, Michael Bitzer, Nisar Malek, Dominik Rath, Hans Bösmüller, Bernard Nieswandt, Meinrad Gawaz, Tamam Bakchoul, Peter Rosenberger

**Author notes:** **Corresponding author:** Tamam Bakchoul, MD Center for Clinical Transfusion Medicine, University Hospital of Tuebingen, Otfried-Mueller-Straße 4/1, 72076 Tuebingen, Germany. Indicates equal contribution (Karina Althaus, Irene Marini). Indicates shared senior authorship (Tamam Bakchoul, Peter Rosenberger).

## Abstract

The pathophysiology of COVID-19 associated thrombosis seems to be multifactorial, involving interplay between cellular and plasmatic elements of the hemostasis. We hypothesized that COVID-19 is accompanied by platelet apoptosis with subsequent alteration of the coagulation system. We investigated depolarization of mitochondrial inner transmembrane potential (ΔΨm), cytosolic calcium (Ca^2+^) concentration, and phosphatidylserine (PS) externalization by flow cytometry. Platelets from intensive care unit (ICU) COVID-19 patients (n=21) showed higher ΔΨm depolarization, cytosolic Ca^2+^ concentration and PS externalization, compared to healthy controls (n=18) and COVID-19 non-ICU patients (n=4). Moreover significant higher cytosolic Ca^2+^ concentration and PS was observed compared to septic ICU control group (ICU control). In ICU control group (n=5; ICU non-COVID-19) cytosolic Ca^2+^ concentration and PS externalization was comparable to healthy control, with an increase ΔΨm depolarization. Sera from ICU COVID-19 13 patients induced significant increase in apoptosis markers (ΔΨm depolarization, cytosolic Ca^2+^ concentration and PS externalization). compared to healthy volunteer and septic ICU control. Interestingly, immunoglobulin G (IgG) fractions from COVID-19 patients induced an Fc gamma receptor IIA dependent platelet apoptosis (ΔΨm depolarization, cytosolic Ca^2+^ concentration and PS externalization). Enhanced PS externalization in platelets from ICU COVID-19 patients was associated with increased sequential organ failure assessment (SOFA) score (r=0.5635) and DDimer (r=0.4473). Most importantly, patients with thrombosis had significantly higher PS externalization compared to those without. The strong correlations between apoptosis markers and increased D-Dimer levels as well as the incidence of thrombosis may indicate that antibody-mediated platelet apoptosis potentially contributes to sustained increased thromboembolic risk in ICU COVID-19 patients.

**Key points:**

1. Severe COVID-19 is associated with increased antibody-mediated platelet apoptosis.
2. Platelet apoptosis in severe COVID-19 is correlated with D-Dimer and higher incidence of thromboembolisms.

## Introduction

Accumulating evidence indicates an association between SARS-CoV-2 associated pneumonia and hypercoagulable state of patients with corona virus disease 2019 (COVID-19) who require intensive care (severe infection)^1^. It becomes be more and more clear that infections with SARS-CoV-2 did not meet the criteria of disseminated intravascular coagualation (DIC) according to the International Society of Thrombosis and Haemostasis (ISTH) and the patterns of coagulation seem to be different to severe infections^2^. Only in patients with severe aggravation of disease an overt DIC occurred^3^. At the same time, however, the incidence of acute pulmonary embolism (PE), deep-vein thrombosis, ischemic stroke, myocardial infarction and/or systemic arterial embolism in COVID-19 patients admitted to the intensive care unit (ICU) is as high as 49%^4^. The pathophysiology of COVID-19 associated-thromboembolic events seems to be complex and multifactorial, involving interplay between cellular and plasmatic elements of the hemostatic system and components of the innate immune response to the infecting pathogen. The phosphatidylserine (PS) externalization in apoptotic platelets, as a substrate, might be an initiation for multiple coagulation factors^5^. So a combination of several activation events initiated by exposure of the endothelium, platelets, and leukocytes to pathogen- and damage-associated molecular patterns might be responsible for the uncontrolled activation of the coagulation system in severely ill COVID-19 patients^6,7^.

To date most clinical reports on COVID-19-associated coagulopathy focused on an increased activation of the plasmatic coagulation system^1-3,7^. Elevated D-Dimer levels were consistently reported, whereas their gradual increase during disease course is particularly associated with disease worsening^8^. Other coagulation abnormalities such as prothrombin time(PT), activated partial thromboplastin time (aPTT) prolongation, together with severe thrombocytopenia have been found to be associated with life-threatening DIC^9^. Platelets have been recently recognized as a mediator of inflammation and sensor of infectious agents through the interaction of surface receptors and pathogens or immune system derivatives^10^. Viral infections elicit the systemic inflammatory response that affects platelets, which become activated upon antigen specific recognition and interaction with white blood cells^11^. While platelet activation plays a critical role in the procoagulatory effect of viral infections^12,13^, platelets derived from patients with HIV as well as secondary dengue virus infection have been shown to have increased upregulation of the intrinsic pathway of apoptosis^14-16^. In addition, it is well known that the survival and lifespan of platelets is finely regulated by the intrinsic or mitochondrial apoptosis pathway^17,18^.

We hypothesized that coagulation disturbances observed in ICU COVID-19 patients are accompanied by platelet apoptosis with subsequent alterations of the coagulation system. In platelets, apoptosis is mediated by the mitochondrial outer membrane permeabilization (MOMP) which is regulated by the members of the Bcl2 protein family that either promote (pro-apoptotic proteins: Bak, Bax and Bim) or inhibit (anti-apoptotic proteins: Bcl-xL and Bcl-w) the apoptosis^19^. The mitochondrial inner membrane potential (ΔΨm) collapse is followed by the efflux of cytochrome c into the cytoplasm which forms a multi-protein complex called apoptosome. The latter triggers the activation of the caspase 9 and the following downstream caspase cascade including caspase 3 and 7. Finally, the phosphatidylserine (PS) externalization on the extracellular membrane represents one of the late stage of the apoptosis pathway^20^. In this study, we found evidence that platelets from severe ICU COVID-19 patients have an upregulation of apoptotic markers. Most importantly, sera and IgG-fractions isolated from COVID-19 patients were able to induce apoptosis in platelets from healthy donors. Furthermore, our data indicate that platelet apoptosis might be associated with thromboembolic complications and increased mortality in severe COVID-19.

## Methods

### Study cohort and evaluation of the clinical data

During the SARS-CoV-2 outbreak in Tuebingen (between March the1^st^ and April the 30^th^ 2020), 27 ICU COVID-19 and non-ICU patients were referred to our laboratory for extended investigations of the coagulations system. Blood samples from healthy donors (n = 18) and septic ICU non-COVID-19 patients (n = 5) were collected to serve as controls.

Electronic medical records were used to collect demographical data, clinical treatments and outcome. To estimate the status of critical illness, the sequential organ failure assessment (SOFA) score system was employed as previously described^21^. Data were independently reviewed by two physicians (K.A. and S.H.). In case of disagreements, a third physician was consulted (M.M.). For more details see supplemental material

### Determination of apoptosis in patients’ platelets

#### Platelet isolation

Platelets were isolated from citrated blood and tested within 3 hours (h*). In brief, after one centrifugation step (120g, 20 minutes [min*] at room temperature [RT], without brake), platelet rich plasma (PRP) was gently separated and used for further analysis.

#### Assessment of the inner-mitochondrial-transmembrane potential (ΔΨm)

To detect changes in the ΔΨm, the tetramethylrhodamine ethyl ester (TMRE) assay kit (Abcam, Cambridge, United Kingdom) was used as previously described, with minor modification^22,23^. In brief, PRP was stained with 10 μM TMRE (30 min* at RT) and directly measured by flow cytometry (FC) (Navios, Beckman-Coulter, USA). The complete depolarization of platelet mitochondrial potential was induced using the uncoupler of mitochondrial oxidative phosphorylation carbonyl cyanide 4-trifluoromethoxy phenylhydrazone (FCCP) (10 μM, 1 h* at 37°C)^16^. Changes in the ΔΨm were determined as ratio of the mean of fluorescence intensity (MFI) signal TMRE from healthy donors’ PRP compared to the MFI signal of platelets from COVID-19 and ICU non-COVID-19 patients.

#### Quantification of cytosolic calcium (Ca^2+^) concentration

Cytosolic Ca^2+^ concentration was determine using Fluo-3/AM (Sigma-Aldrich, St. Louis, USA) as previously described^24^, with minor modifications. In brief, PRP were incubated with Fluo3/AM (3 μM) in TRIS (trisaminomethane) buffer (10 mM TRIS, 0.9% NaCl, 1 mM CaCl_2_, pH 7.4) for 30 min* at 37°C. After washing once with TRIS buffer, cells were resuspended in TRIS containing 1 mM CaCl_2_. Ca^2+^-dependent fluorescence intensity of samples was measured with an excitation wavelength of 488 nm and an emission wavelength of 530 nm by FC. PRP treated with ionomycin (2 μM, 30 min* at 37°C, [Sigma-Aldrich, St. Louis, USA]) served as positive control. Test results were normalized to the mean MFI of healthy donors tested in parallel.

#### Detection of phosphatidylserine externalization

PRP was stained with Annexin V-FITC (Immunotools, Friesoyhte Germany) and CD41-PE-Cy5 (Beckman Coulter, Brea, USA) in TRIS buffer (30 min* at RT) and directly analyzed by FC, as previously described^25^. Cells incubated with 60 μM ionomycin (30 min* at RT) served as positive control. Test results were determined as fold increase of the percentage of PS positive events in PRP from COVID-19 or ICU non-COVID-19 patients compared to platelets from healthy donors tested in parallel.

### Assessment of antibody-mediated apoptosis

#### Sera and IgG incubation with washed platelets

To exclude unspecific effects like the activation of platelets via complement or non-specific immune complexes, all sera were heat-inactivated (56°C for 30 min*), followed by a sharp centrifugation step at 5,000g. The supernatant was collected.

All experiments involving patients’ sera were performed after incubation of 5 μL serum with 25 μL washed platelets (7.5×10^6^, see supplemental material for more information on the preparation of washed platelets) for 1.5 h* under rotating conditions at RT. Afterwards, samples were washed once (7 min*, 650g, RT, without brake) and gently resuspended in 75 μL of phosphate-buffered saline (PBS, Biochrom, Berlin, Germany).

When indicated, immunoglobulin G (IgG) fractions were isolated from serum (for more details see supplemental material).

#### Detection of ΔΨm changes induced by patients’ sera

Washed platelets were stained with 10 μM TMRE (30 min* at RT) and directly measured by FC. As positive control, cells were preincubated with FCCP (10 μM for 30 min* at 37°C). Serum-mediated apoptosis was quantified as ratio of ΔΨm depolarization comparing the MFI of washed platelets incubated with serum from healthy donors with sera from (non-ICU and ICU) COVID-19 or ICU non-COVID-19 patients. Test results were normalized to the mean MFI of patients’ sera compared to sera from healthy donors tested in parallel.

#### Quantification of cytosolic Ca^2+^ concentration after patients’ sera incubation

Upon incubation with patients’ sera washed platelets were incubated with Fluo3/AM (3 μM) in TRIS buffer (30 min* at 37°C). After washing once with TRIS buffer, cells were resuspended in TRIS supplemented with 1 mM CaCl_2_. Subsequently, Ca^2+^-dependent fluorescence intensity of samples was measured by FC. Ionomycin (5 μM, 15 min* at RT) was used as positive control. Test results were normalized to the mean MFI of patients’ sera compared to sera from healthy donors tested in parallel.

#### Assessment of phosphatidylserine externalization induced by patients’ sera

Washed platelet (1×10^6^) were resuspended into Hank’s balanced salt solution (137 mM NaCl, 1.25 mM CaCl_2_, 5.5 mM glucose [Carl-Roth, Karlsruhe, Germany]), stained with Annexin V-FITC (Immunotools, Friesoythe Germany) and directly analyzed by FC. As positive control washed platelets were incubated with ionomycin (5μM, 15 min* at RT). Test results were determined as fold increase of the percentage of PS positive events in platelets upon incubation with patients’ sera compared to cells incubated with healthy donors tested in parallel.

### Western blot analysis

Protein levels of cleaved-caspase 9 were determined by western blot. In brief, washed platelet were centrifuged (5 min*, 700g at 4°C) and the pellet was resuspended in RIPA lysis buffer (ThermoFisher Scientific, Paisley, UK). The proteins were separated by electrophoresis using 12% SDS-PAGE. See the supplemental material for further details.

### Ethics Statement

The study was conducted in accordance with the declaration of Helsinki. Written informed consent was obtained from all volunteers prior to any study-related procedure. All tests were performed with rest material from routine testing. The study protocol of patient material was approved by the Institutional Review Board of the University of Tuebingen.

### Statistical analyses

The statistical analysis was performed using GraphPad Prism, Version 7.0 (GraphPad, La Jolla, USA). Because potential daily variations in FC measurements might result in bias in data analysis, test results were normalized to two healthy donors tested in parallel at the same time point (raw data are available in the supplemental data). For further information see the supplemental material.

### Data sharing statement

Data may be requested for academic collaboration from the corresponding author.

## Results

### Study cohort

Blood samples were collected from 27 consecutive COVID-19 patients who were admitted to our hospital with severe acute respiratory distress symptoms (ARDS) requiring intensive care (ICU, n = 23) and hospitalized patients (non-ICU n = 4) without ARDS. Two blood samples were excluded due to insufficient material. In total, 21 ICU COVID-19 patients were enrolled in the study, of whom 18/21 (86%) patients were male, Table 1. The mean age was 60 years (range: 29–88 years, Table 1). 15/21 (71%) patients had known risk factors for severe COVID-19 infection as described previously^26^, including hypertension in 14/21 (67%), obesity in 4/21 (19%), coronary artery disease 4/21 (19%) and diabetes mellitus 5/21 (24%). 4 patients had only minor symptoms of COVID-19 and did not require admission to the ICU. Samples from patients on ICU with sepsis were used as a control group. Detailed information on the clinical manifestations of the 25 (non-ICU, n = 4 and ICU, n = 21) COVID-19 patients and the 5 ICU non-COVID-19 patients (ICU control) are available in the supplemental data (Supplemental Table 1).

**Table 1.** Baseline parameters of 21 intensive care unit (ICU) COVID-19 patients

|  | Total<br>(n=21) | Survivors<br>(n=14) | Non-survivors<br>(n = 7) | *p-value* |
| --- | --- | --- | --- | --- |
| Age, years (mean ± SD) | 60 (±17) | 58 (±18) | 63 (±16) | 0.359 |
| Male, n (%) | 18 (85.7) | 11 (78.6) | 7 (100) | 0.186 |
| Body mass index (mean ± SD) | 28.6 (±5) | 28 (±5) | 29 (±4) | 0.543 |
| **Cardiovascular risk factors, n (%)** |  |  |  |  |
| Arterial hypertension | 14 (66.7) | 9 (64.3) | 5 (71.4) | 0.743 |
| Dyslipidemia | 5 (23.8) | 3 (21.4) | 2 (28.6) | 0.717 |
| Diabetes mellitus | 5 (23.8) | 4 (28.6) | 1 (14.3) | 0.469 |
| Current smokers | 0 (0.0) | 0 (0.0) | 0 (0.0) | NA |
| Obesity* | 4 (19.0) | 3 (21.4) | 1 (14.3) | 0.807 |
| Atrial fibrillation | 4 (19.0) | 4 (28.6) | 0 (0.0) | 0.116 |
| Known CAD | 4 (19.0) | 2 (14.3) | 2 (28.6) | 0.597 |
| Chronic kidney disease | 0 (0.0) | 0 (0.0) | 0 (0.0) | NA |

| **Echocardiography** |  |  |  |  |
| --- | --- | --- | --- | --- |
| Left ventricular function, % (mean ± SD) | 56 (±6) | 57 (±7) | 54 (±6) | 0.516 |
| RV pressure, mmHg (mean ± SD) | 26 (±3) | 23 (±6) | 33 (±15) | 0.401 |
| **Admission laboratory, median (25th percentile - 75th percentile)** |  |  |  |  |
| Leucocytes, 1000/μL | 6.5 (4.2-9.7) | 6.7 (4.3 - 11.0) | 5.6 (3.0 - 6.8) | 0.233 |
| Lymphocytes, 1000/μL | 0.6 (0.3-1.0) | 0.7 (0.5 - 1.1) | 0.3 (0.3 - 0.8) | **0.044** |
| PLT, x10<sup>9</sup>/L | 179 (152-262) | 165 (87-218) | 182 (154-336) | 0.154 |
| INR | 1.0 (1.0-1.1) | 1.0 (1.0-1.1) | 1.1 (1.0-1.1) | 0.474 |
| aPTT, s* | 26 (24-30) | 25 (22-30) | 28 (24-31) | 0.2609 |
| Fibrinogen, mg/dL | 574 (482-689) | 613 (495-730) | 494 (463-574) | 0.2242 |
| D-Dimer, μg/dL | 1.7 (1.0-5.5) | 1.8 (1.0 - 4.4) | 1.2 (0.9 - 18.0) | 1.000 |
| GFR, mL/m<sup>2</sup> | 73.9<br>(42.4-93.2) | 65.8<br>(43.1 - 87.8) | 81.6<br>(26.7 - 139.4) | 0.296 |
| Creatinin, mg/dL | 1.0 (0.8-1.7) | 1.1 (0.9 - 1.7) | 0.9 (0.6 - 2.5) | 0.331 |
| C-reactive protein, mg/dL | 17. Jan<br>(11.8-27.6) | 15.0<br>(12.1 - 19.1) | 28. Mrz<br>(7.7 - 29.5) | 0.136 |
| Procalcitonin, ng/mL | 0.7<br>(0.2-1.8) | 0.7<br>(0.2 - 2.8) | 0.2<br>(0.2 - 2.0) | 0.823 |
| Troponin I, ng/dL | 26 (13.8-111.3) | 29 (16 - 210) | 22 (6 - 34) | 0.284 |
| NT pro-BNP, ng/L | 749<br>(385-3148) | 1169<br>(239 - 4543) | 537<br>(385 - 2324) | 0.661 |
| CK, U/L | 312<br>(181-1166) | 300<br>(159 - 1416) | 414<br>(280 - 1057) | 0.602 |
| AST, U/L | 62 (45-118) | 63 (43 - 134) | 61 (47 - 108) | 0.881 |
| ALT, U/L | 45 (28-76) | 46 (29 - 68) | 34 (19 - 88) | 0.455 |
| LDH, U/L | 415 (375-498) | 426 (382 - 504) | 394 (312 - 497) | 0.502 |
| **Medication at admission, n (%)** |  |  |  |  |
| Oral anticoagulation | 1 (4.8) | 1 (7.1) | 0 (0.0) | 0.424 |
| ACEi/ARB | 9 (42.9) | 5 (35.7) | 4 (57.1) | 0.515 |
| Aldosterone inhibitors | 2 (9.5) | 0 (0.0) | 2 (28.6) | 0.051 |
| Diuretics | 2 (9.5) | 1 (7.1) | 1 (14.3) | 0.696 |
| Calcium channel blockers | 3 (14.3) | 2 (14.3) | 1 (14.3) | 0.869 |
| Beta blockers | 7 (33.3) | 3 (21.4) | 4 (57.1) | 0.152 |
| Statins | 5 (23.8) | 2 (14.3) | 3 (42.9) | 0.210 |
| ASA | 5 (23.8) | 2 (14.3) | 3 (42.9) | 0.210 |
| P2Y12 blockers | 0 (0.0) | 0 (0.0) | 0 (0.0) | NA |

ACEi indicates angiotensin-converting enzyme inhibitors; ALT, Alanine Aminotransferase; aPTT, activated partial thromboplastin time; ARB, Angiotensin II receptor blockers; ASA, acetylsalicylic acid; AST, Aspartate aminotransferase; CAD, cardiac artery disease; CK, creatine kinase; GFR, Glomerular Filtration Rate; INR, international normalized ratio; LDH, Lactate dehydrogenase; NT pro BNP, N-terminal pro b-type natriuretic peptide; PLT, platelet; RV, right ventricular; s\*, seconds; SD, standard deviation; \*Obesity (body mass index, BMI>30).

On the ICU, all patients received controlled respiratory support and six of them had additionally venous-venous extracorporeal membrane oxygenation (vv-ECMO). Anticoagulation with heparin was administrated at prophylactic doses (400 IE/h) in 12/21 (57%) patients or at therapeutic doses (2 to 3 fold aPTT) in 9/21 (43%) patients, depending on patient’s risk factors for thrombosis. 8/21 (38%) patients had additional antiplatelet therapy with aspirin. At the day of material submission, patients had a median SOFA score of 8 (range: 2–17), median platelet counts of 151×10^9^/L (range: 35–500×10^9^/L), median aPTT 39 seconds (s*) (range: 20–98 s*), median international normalized ratio (INR) of 1.0 (range: 0.9–1.5), median D-Dimer 4 μg/mL (range: 1–42 μg/mL) and median fibrinogen 551 mg/dL (range: 273–961 mg/dL), Supplemental Table 1.

During the 30-day follow-up, 7/21 (33%) deceased, 13/21 (62%) patients developed thrombocytopenia (range platelet count: 9–149×10^9^/L) and 12/21 (57%) patients had at least one thromboembolic complication including kidney, spleen and liver infarction (n = 7), catheter associated thrombosis (n = 2), clotting of extracorporeal circulation system (n = 1), pulmonary embolism (n = 1), cerebral infarction (n = 1) and myocardial infarction (n = 1), Supplemental Table 1.

### Severe COVID-19 infection is associated with platelet apoptosis

To evaluate the mitochondrial function in platelets from COVID-19 patients, the mitochondrial ΔΨm, was assessed. As shown in Figure 1A, significantly increased ΔΨm depolarization was found in platelets isolated from ICU COVID-19 patients compared to healthy donors (1.385 ±0.071 vs. 0,997 ±0,050, p = 0.0001) as well as to non-ICU COVID-19 patients (1.385 ±0.071 vs. 0.851 ±0.096, p = 0.0046, Figure 1A; Supplemental figure 1A). Of note, a comparable ΔΨm depolarization was observed between ICU COVID-19 and ICU non-COVID-19 patients (1.385 ±0.071 vs.1.369 ±0.112, p = 0.921, Figure 1A; Supplemental figure 1A).

**Figure 1.**
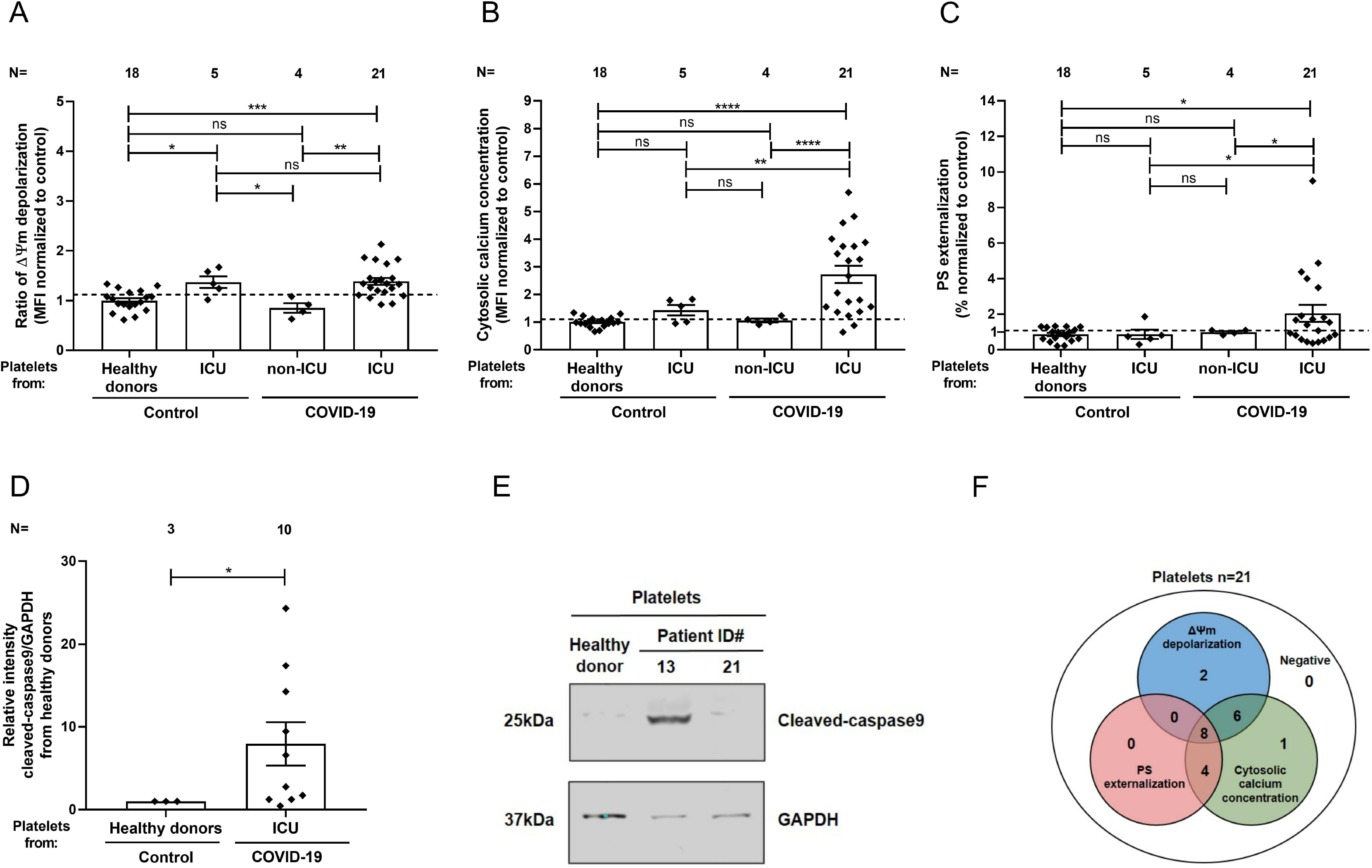
Platelet apoptosis in COVID-19 patients. Changes in apoptosis pathways were analyzed by assessing the depolarization of the mitochondrial inner transmembrane potential (ΔΨ m) (A), cytosolic calcium concentration (B), and phosphatidylserine (PS) externalization (C) in platelets from intensive care unit (ICU) COVID-19 or non-ICU COVID-19 patients as well as ICU non-COVID-19 patients (control group) and healthy donors, respectively. (D) Quantification of cleaved-caspase 9 level in platelet from ICU COVID-19 patients normalized to healthy donors. (E) Representative western blot showing GAPDH and cleaved-caspase 9 proteins in platelet from ICU COVID-19 patients. Proteins bands were detected with the infrared imaging system (Odyssey, LI.COR^®^, Lincoln, USA). (F) Diagram indicating the number of ICU COVID-19 patients positive for each apoptotic parameter: ΔΨm depolarization, cytosolic calcium concentration and PS externalization. Data are presented as mean ± standard error mean (SEM) of the measured fold increase compared to control, not significant, *p< 0.05, **p< 0.01, ***p< 0.001 and ****p< 0.0001. The number of patients and healthy donors tested is reported in each graphic. Dot lines represent the cutoffs determined from healthy donors as mean of fold increase (FI) + 2× SEM.

Next, we determined the cytosolic Ca^2+^ concentration in platelets from COVID-19 patients using the intracellular dye Fluo-3. Platelets from ICU COVID-19 patients showed a significant increase in the cytosolic Ca^2+^ concentration compared to healthy donors (2.729 ±0.314 vs. 1.007 ±0.043, p< 0.0001), to non-ICU COVID-19 patients (2.729 ±0.314 vs. 1.060 ±0.069, p< 0.0001, as well as to ICU non-COVID-19 patients (2.729 ±0.314 vs. 1.432 ±0.190, p = 0.0018, Figure 1B; Supplemental Figure 1B) was found.

To further confirm the potential involvement of platelet apoptosis in ICU COVID-19 patients, we evaluated surface PS externalization on platelets. The latter, was significantly higher on platelets from ICU COVID-19 patients compared to healthy volunteers (2.051 ±0.476 vs. 0.865 ±0.088, p = 0.0290) as well as to non-ICU COVID-19 patients (2.051 ±0.476 vs. 0.993 ±0.064, p = 0.0392, Figure 1C; Supplemental Figure 1C) as well as ICU non-COVID-19 patients (2.051 ±0.476 vs. 0.869 ±0.263, p = 0.0404, Figure 1C; Supplemental Figure 1C). Moreover, western blot analyses showed increased level of cleaved-caspase 9 levels in platelets from ICU COVID-19 patients compared to healthy donors (7.94 ±2.61 vs. 1.00 ±0.00, p = 0.0260; Figure 1D-E). In association with mitochondrial dysfunction and increased cytosolic Ca^2+^ concentration, these data indicate an activation of the intrinsic pathway of apoptosis in platelets during COVID-19 infection. Overall, as shown in Figure 1F, platelets from 16/21 (76%) patients showed ΔΨm depolarization, 19/21 (90%) had increased cytosolic Ca^2+^ concentration and 12/21 (57%) patients reveled enhanced PS externalization Figure 1F.

### Association of apoptosis markers with laboratory parameters and clinical outcomes

To draw conclusion on potential clinical impact of our findings, we limited the analysis of the correlations apoptosis markers to platelet count, SOFA score and D-Dimer levels at the days of blood sampling. We found that platelet count is negatively correlated with PS externalization (r = –0.6177, p = 0.0028, Figure 2A) and with cytosolic Ca^2+^ concentration (r = –0.5666; p = 0.0299, Figure 2B). Strong positive correlation was also observed for PS externalization with D-Dimer (r = 0.4473; p = 0.0420; Figure 2C) as well as with the SOFA score (r = 0.5635; p = 0.0078, Figure 2D).

**Figure 2.**
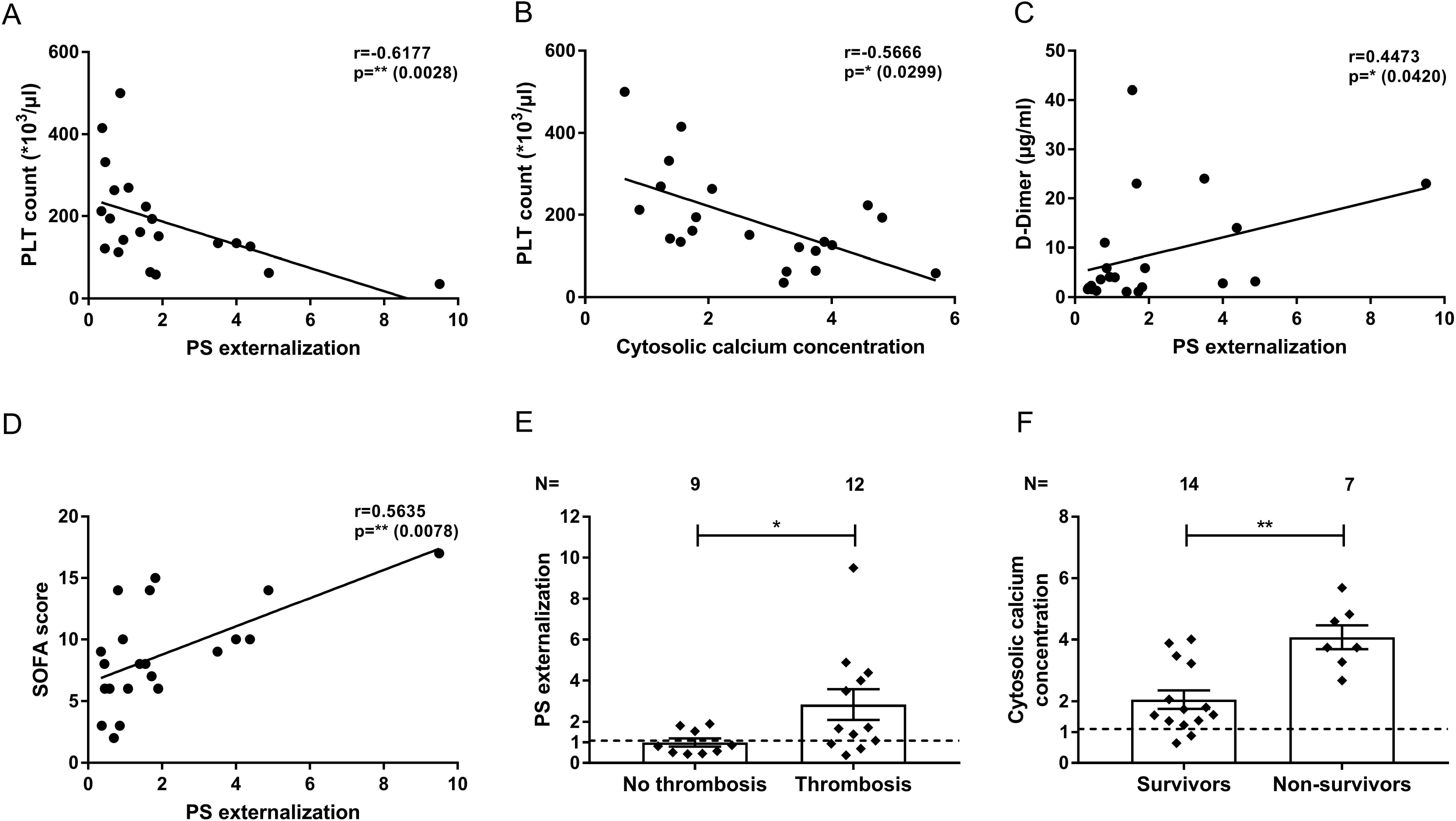
Association between platelet apoptosis and clinical biomarkers, thromboembolic complications and mortality. The correlations between platelet apoptosis parameters and PLT count as well as D-Dimer and SOFA score measured at the same day of platelet testing were assessed. An association was observed between PLT count and phosphatidylserine (PS) externalization (A) as well as cytosolic calcium concentration (B), respectively. Moreover, a significant correlation was detected for D-Dimer and PS externalization (C). The clinical relevance of PS externalization was assessed using the SOFA score and revealed significant positive correlation (D). Pearson’s correlation coefficients were calculated and are shown in the panels. The phosphatidylserine (PS) externalization (E) and the cytosolic calcium concentration (F) were determined and compared between intensive care unit (ICU) COVID-19 patients depending on the incidence of thromboembolic complications and mortality, respectively. Data are presented as mean of the measured fold increase compared to control, not significant, *p< 0.05, **p< 0.01, ***p< 0.001 and ****p< 0.0001. The number of patients and healthy donors tested is reported in each graphic. Dot lines represent the cutoffs determined from healthy donors as mean of fold increase (FI) + 2x standard error mean (SEM).

Next, we analyzed the markers of apoptosis in relation to the incidence of thromboembolic complications and mortality during the 30-day follow-up. We found that ICU COVID-19 patients with thromboembolic complications had significantly higher levels of PS externalization compared to those without thrombosis (2.85 ±0.75 vs. 0.99 ±0.20, p = 0.0340, Figure 2E). In addition, cytosolic Ca^2+^ concentration was significantly higher in non-survivors compared to survivors (4.07 ±0.39 vs. 2.05 ±0.30, p = 0.0012, Figure 2F).

### Sera from ICU COVID-19 patients cause IgG-mediated platelet apoptosis via cross-linking Fc gamma receptor IIA

To explore the mechanism of platelet apoptosis in COVID-19, patients’ sera were incubated with washed platelets from healthy donors. As shown in Figure 3A, COVID-19 sera induced a significantly higher ΔΨm depolarization compared to sera from healthy donors (1.52 ±0.117 vs. 0.958 ±0.082, p = 0.0086) as well as to sera from ICU non-COVID-19 patients (1.52 ±0.117 vs. 1.099 ±0.057, p = 0.0036, Figure 3A; Supplemental Figure 2A). Cytosolic Ca^2+^ concentration was also significantly increased compared to healthy volunteers (1.372 ±0.074 vs. 0.984 ±0.055, p = 0.0036, Figure 3B; Supplemental Figure 2B) and to ICU non-COVID-19 patients (1.372 ±0.074 vs. 1.086 ±0.066, p = 0.0113, Figure 3B; Supplemental Figure 2B). Similarly, enhanced PS externalization was found in platelet upon incubation with ICU COVID-19 sera compared to healthy donors (1.624 ± 0.126 vs. 0.969 ±0.100, p = 0.0051, Figure 3C; Supplemental Figure 2C) as well as to ICU non-COVID-19 sera (1.624 ± 0.126 vs. 0.953 ±0.169, p = 0.0214, Figure 3C; Supplemental Figure 2C). In addition, significantly higher levels of cleaved-caspase 9 were found after incubation with sera from COVID-19 compared to healthy control (3.40 ±0.65 vs. 1.00 ±0.00, p = 0.0030; Figure 3D-E). In the whole cohort, sera from 13/21 (62%) patients induced enhanced ΔΨm depolarization, 8/19 (42%) patients increased cytosolic Ca^2+^ concentration and 13/21 (62%) patient^’^s sera mediated increased PS externalization on platelets of healthy donors (Figure 3F).

**Figure 3.**
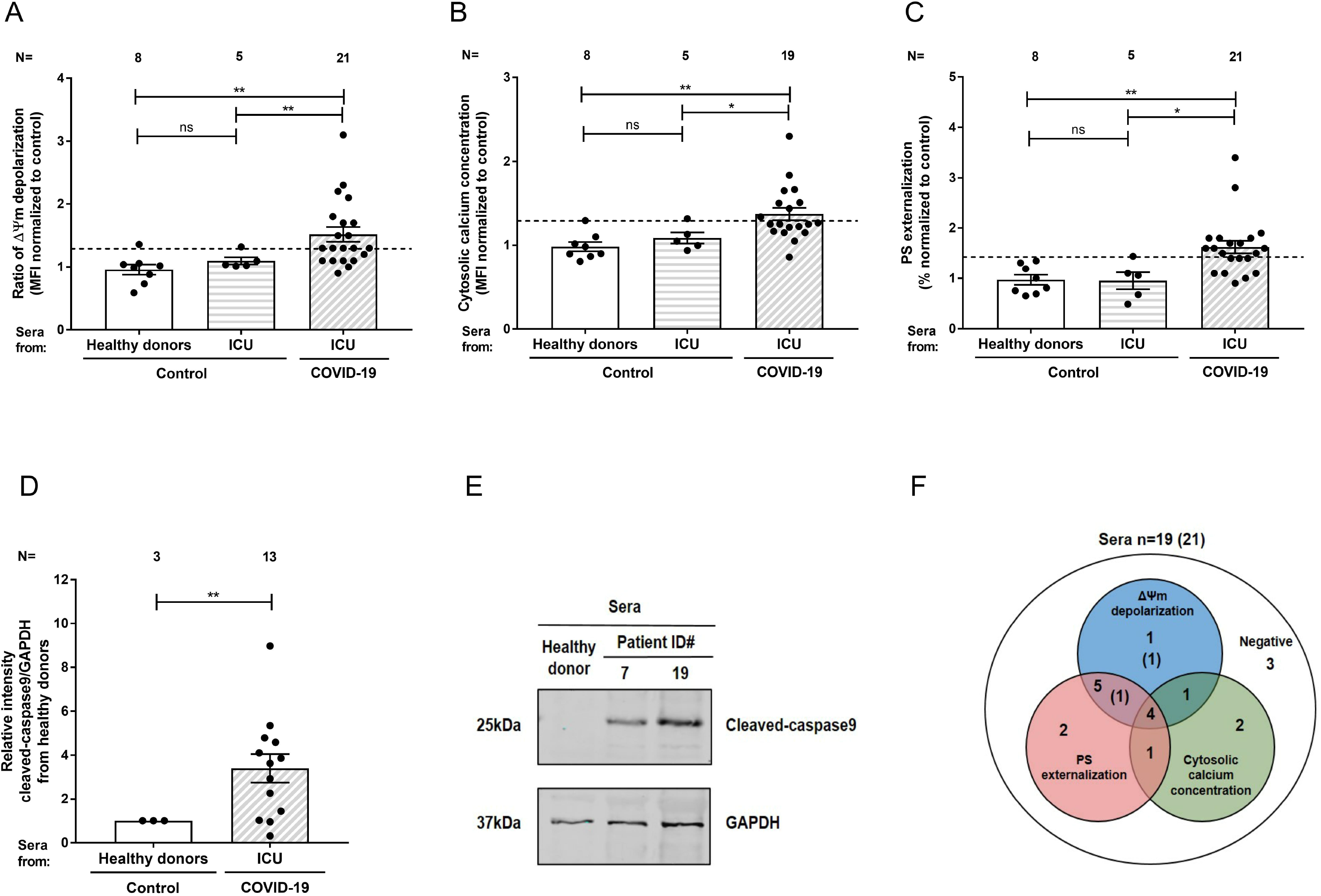
Impact of sera from intensive care unit (ICU) COVID-19 patients on platelet apoptosis. Changes in apoptosis pathways induced by sera from ICU COVID-19 patients, ICU non-COVID-19 patients (control group) and healthy donors were investigated by assessing the depolarization of the mitochondrial inner transmembrane potential (ΔΨ m) (A), cytosolic calcium concentration (B, n = 19 due to the lack of biomaterial for 2 patients), and PS externalization (C). (D) Quantification of cleaved-caspase 9 level in platelet from healthy donors after incubation with patients’ sera, normalized to sera from healthy donors. (E) Representative western blot of GAPDH and cleaved-caspase 9 proteins. Proteins’ bands were detected with the infrared imaging system (Odyssey, LI.COR^®^, Lincoln, USA). (F) Diagram indicating the number of ICU COVID-19 patients positive for each apoptotic parameter: ΔΨm depolarization, cytosolic calcium concentration and PS externalization. In round brackets are reported the two sera tested only for: ΔΨm depolarization and PS externalization. Data are presented as mean ± standard error mean (SEM) of the measured fold increase compared to control, not significant, *p< 0.05, **p< 0.01, ***p< 0.001 and ****p< 0.0001. The number of sera tested is reported in each graphic. Dot lines represent the cutoffs determined testing sera from healthy donors as mean of fold increase (FI) + 2× SEM.

In order to further dissect the pathways of the antibody-mediated platelet apoptosis, IgG-fractions were isolated from apoptosis-inducing sera tested in the absence or presence of an FcγRIIA-blocking mAb (IV.3). Interestingly, IgG fractions from COVID-19 sera induced significant upregulation in all three apoptosis markers, ΔΨ m depolarization (1.54 ±0.14 vs. 1.00 ±0.00, p = 0.0051, Figure 4A), cytosolic Ca^2+^ concentration (1.36 ±0.05 vs. 1.00 ±0.00, p< 0.0001, Figure 4B) and PS externalization (1.70 ±0.27 vs. 1.00 ±0.00, p = 0.0330, Figure 4C) compared to healthy donors. In contrast, a significant reduction of platelet apoptosis was observed after blocking platelet FcγRIIA: ΔΨm depolarization (2.23 ±0.25 vs. 1.22 ±0.12, p = 0.0216, Figure 4D), cytosolic Ca^2+^ concentration (1.80 ±0.15 vs. 0.58 ±0.04, p = 0.0015, Figure 4E) and PS externalization on platelets(9.59 ±1.52 vs. 2.12 ±0.20, p = 0.0371, Figure 4F), respectively. Of note, protein staining of the isolated IgG showed no hint of IgG aggregates or immune complexes (data not shown).

**Figure 4.**
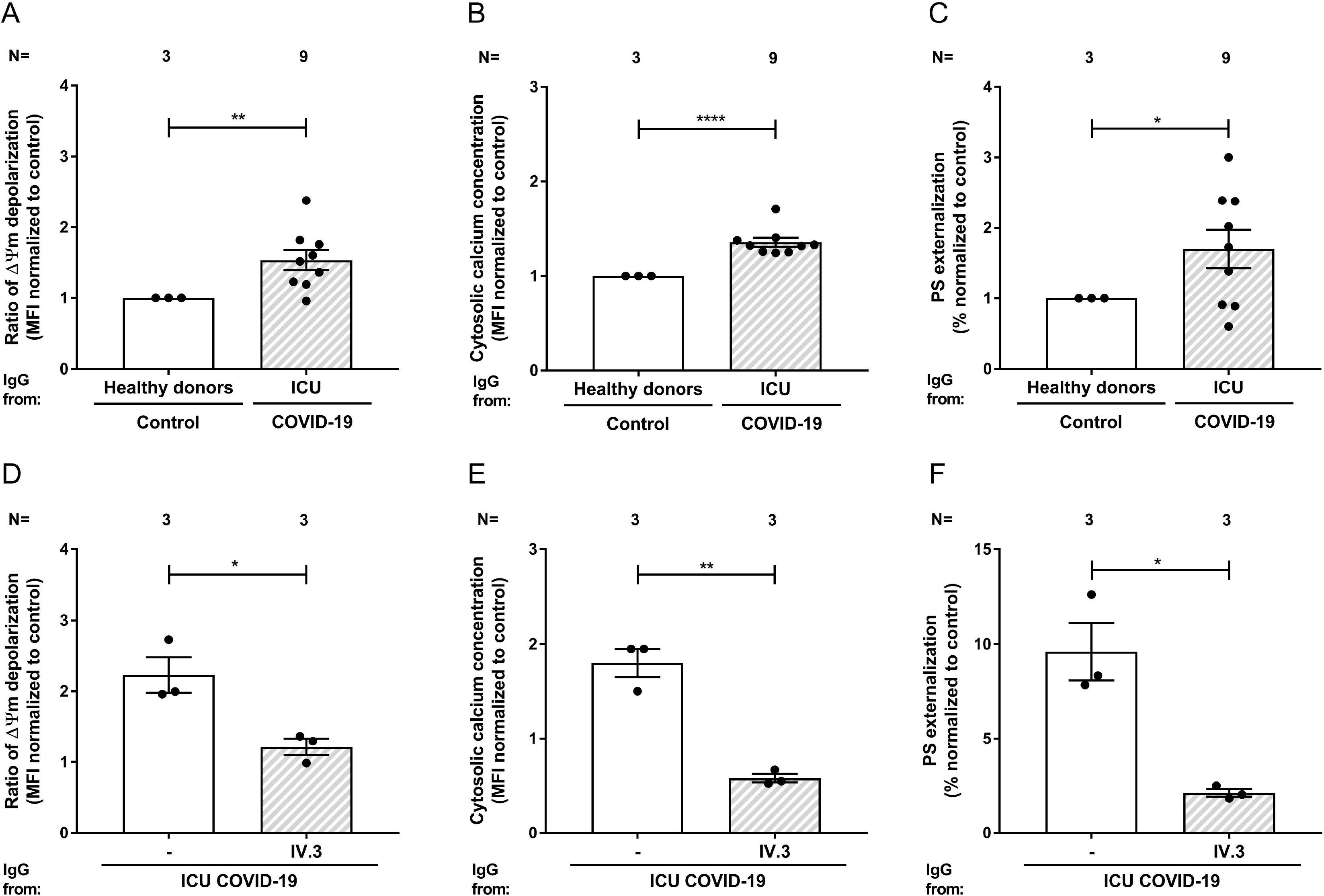
IgG fraction from intensive care unit (ICU) COVID-19 patients induce platelet apoptosis via cross linking Fc gamma RIIA. Changes in apoptosis pathways induced by IgG fraction from ICU COVID-19 patients or healthy donors were analyzed by assessing the depolarization of the mitochondrial inner transmembrane potential (ΔΨ m) (A), cytosolic calcium concentration (B), and phosphatidylserine (PS) externalization (C) in platelets from three different healthy donors. The same assays were performed in the presence of the IV.3 monoclonal antibody (mAb) in order to block the FcγRIIA signaling (D, E and F), respectively. Each dot represents two experiments with platelets from two different donors. Data are presented as mean ± standard error mean (SEM) of the measured fold increase compared to control, not significant, *p< 0.05, **p< 0.01, ***p< 0.001 and ****p< 0.0001. The number of sera tested is reported in each graphic.

## Discussion

COVID-19-infected individuals have a heightened risk for developing thromboembolic complications and a link has been suggested between low platelet count and severity of disease and mortality^27-29^. Here, we show that severe COVID-19 is associated with antibody-mediated upregulation of platelet apoptosis. In addition, we found a correlation between platelet apoptosis markers and SOFA score, plasma levels of D-Dimer as well as the incidence of thromboembolic complications in severe COVID-19 patients. These data indicate that platelet apoptosis may contribute to sustained inflammation and increased thromboembolic risk in COVID-19 patients and could potentially present a potential therapeutic target.

A subpopulation with severe COVID-19 in our study had an increase in platelet apoptosis markers. The exact mechanism of COVID-19 induced platelet apoptosis has not been studied so far. Our data show depolarization of mitochondrial inner transmembrane potential in a subgroup and enhanced cytosolic Ca^2+^ concentration in most of the patients with severe (ICU) COVID-19. Together with the increased PS externalization and cleavage of caspase 9, these results suggest that platelet apoptosis in severe COVID-19 is activated via the intrinsic pathway. Apoptosis is also described for septic patients^30^. But apoptosis seem to be different from apoptosis in septic patients. Notably, we could find increased ΔΨm depolarization in platelets of septic patients, but we could not show any increase in cytosolic Ca^2+^ concentration or in PS externalization patient platelets in the ICU control group. After incubation of serum of healthy donors we see no effect of septic patients. As a difference to platelet apoptosis in septic patients, there might be a serum factor for apoptosis in COVID-19 patients.

While, no enhanced apoptosis was observed in patients platelets or in healthy platelets after incubation of sera isolated from ICU non-COVID-19 control patients suggesting that the activation of the apoptotic pathway may be a specific effect induced by the COVID-19 infection through a serum component. But it remain unclear whether apoptosis alone is sufficient to support thromboembolic status in these patients or if it is linked to platelet activation as well, as increased cytosolic Ca^2+^ is also a finding in increased activated platelets. In apoptotic platelets lifetime is shortened and platelet function is also reduced. In our cohort severe thrombocytopenia was not common among COVID-19 patients, which was also described for the SARS-CoV-1 in 2008^31^. In fact, SARS-CoV-1, which caused the epidemic outbreak in 2003, has also been previously shown to induce apoptosis in Vero cells in a virus replication-dependent manner^32^, despite missing thrombocytopenia. Downregulation of Bcl-2, the activation of effector caspase 3, as well as the upregulation of the pro-apoptotic protein Bax were detected, suggesting the involvement of the caspase family and the activation of the mitochondrial signaling pathway. In fact, our data showed also that platelets from severe COVID-19 patients have higher levels of cleaved-caspase 9.

Prolonged exposure to higher concentrations of platelet agonists has been shown to induce the collapse of mitochondrial membrane potential and subsequent platelet apoptosis^33^. Enhanced platelet activation leading to platelet deposition in damaged pulmonary blood vessels has been demonstrated recently for COVID-19^34^. To exclude potential contribution of platelet hyperactive status in our cohort, we assessed the impact of patients’ sera on platelet apoptosis. Recent data showed that autoantibodies from patients with autoimmune thrombocytopenia (ITP) are able to induce platelet apoptosis^35^. The incubation of sera as well as from IgG fractions from severe COVID-19 patients with platelets from healthy donors induced significant changes in apoptosis markers including ΔΨm depolarization, increased cytosolic Ca^2+^ concentration, caspase 9 cleavage, and finally PS externalization. More importantly, the antibody-mediated platelet apoptosis was inhibited by blocking the FcγRIIA receptors using a specifc mAb. It remains unclear, weather it is a direct IgG-virus complex interaction like previously described for the influenza virus H1N1 infection^36^ and dengue fever or an indirect effect after binding to platelet non-specific targets.

Nevertheless, our data indicate that severe COVID-19 is associated with antibody-mediated activation of the intrinsic pathway via crosslinking FcγRIIA receptors by IgG antibodies against tobe-identified target antigen(s). These findings might offer new therapeutic options like blockade of the FcγRIIA receptor sginaling by tyrosinkinase inhibitors, which have suggested to have a potential use to prevent platelet activation in heparin-induced thrombocytopenia (HIT)^37^.

The clinical relevance of platelet apoptosis is supported by the significant negative correlation between PS levels on platelet surface with platelet count, the positive correlation with DDimer plasma levels, and most importantly with the SOFA score. Meanwhile, it is well established that COVID-19 is associated with increased risk for thromboembolic complications. During the 30-day follow-up, 57% of our patients had thromboembolic complications. Platelet from ICU COVID-19 patients with thromboembolic events showed significantly increased externalization of the apoptosis marker PS compared to those with no thrombosis. This finding suggests a link between thromboembolism and platelet apoptosis in COVID-19 as has been suggested for acute pulmonary embolism^38^.

Our study is subjected to limitations. First, as an observational study, we cannot conclude that the reported associations between platelet markers and laboratory parameters as well as clinical outcomes are causal or specific for infection with SARS-CoV-2. Second, we cannot exclude the possibility of remaining residual confounding or unmeasured potential confounders. Third, the low number of patients does not enable a final and robust multivariate statistical analysis. In addition, we observed a small difference in age between ICU COVID-19 patients and controls. However, age of the patient is not classically associated, and little is known on the impact of age on platelet apoptosis.

It should also be emphasized that it is not clear whether the platelet apoptosis described here are drivers of disease severity or a mere consequence of hypercoagulation in severe (ICU) COVID-19 patients. Indeed, the definitive pathophysiology of COVID-19 and answering questions of causality will likely await the development of model systems for the disease. We hope though that our findings present another piece of the puzzle and will motivate further research into the role of platelet apoptosis in COVID-19 associated thromboembolic complications.

Data presented in this study may build a basis for future studies to dissect platelet-mediated pathological mechanisms involved in the progression of COVID-19 and a larger and multi-center study is warranted to investigate the predictive power of platelet apoptosis markers in well-phenotyped longitudinal cohorts. Furthermore, given the dichotomy we found here between PS externalization and thromboembolic complications in our cohort, it is intriguing, especially in the absence of antiviral and immunologic solutions to the current pandemic, to explore the benefit of anti-apoptotic therapies. In particular, different caspase inhibitors that are currently tested in clinical trials (Emricasan, identification number: NCT03205345 and IDN-6556, identification number: NCT00080236) may represent a part of personalized strategy for some individuals affected by COVID-19 who are at high risk for progression to cardiovascular complications.

In summary, we introduce the first description of platelet apoptosis in severe (ICU) COVID-19 patients, suggesting a strong link between platelet apoptosis and thrombosis. These data may indicate platelet apoptosis as a potential target of COVID-19 treatment as has been suggested for ARDS^39^ and other platelet disorders^35,40^.

## Data Availability

Data may be requested for academic collaboration from the corresponding author.

## Acknowledgments

This work was supported by grants from the German Research Foundation and from the Herzstiftung to T.B. (BA5158/4 and TSG-Study), DFG CRC/TR 240 “Platelets-Molecular, cellular and systemic functions in health and disease” (Project # 374031971) TP A03 (to H.S.), TP B01 (to M.G.), TP B07 (to P.R. and B.N.) and TÜFF-Gleichstellungsförderung to K.A. (2563–0–0). We thank our students Umut Tasdelen, Andreas Witzemann and Jonas Funk for their great contribution as well as Wissam Abou Khalel, Karoline Weich and Flavianna Rigoni for their excellent technical support.

## Authorship contributions

K.A., T.B. and P.R. designed the study. H.H., M.M., M.B., N.M., M.G. and P.R. were responsible for the treatment of the patients. K.A., S.H., D.R. and M.M. collected and analyzed the clinical data. K.A., I.M., J.Z. and L.P. performed the experiments. I.M., K.A., T.B. and P.R. collected the data, H.B. provided pathological finding on thrombosis. K.A., I.M., J.Z., L.P., H.S., B.N., T.B. and P.R. analyzed the data, interpreted the results and wrote the manuscript. All authors read and approved the manuscript.

